# First report of tocilizumab use in a cohort of Latin American patients hospitalized for severe COVID-19 pneumonia

**DOI:** 10.1101/2020.08.12.20173104

**Authors:** Omar Valenzuela, Sebastián Ibáñez, María Poli, Patricia Roessler, Mabel Aylwin, Gigia Roizen, Mirentxu Iruretagoyena, Vivianne Agar, Javiera Donoso, Margarita Fierro, José Montes

## Abstract

**Introduction/objectives:** An interleukin-6 inhibition strategy could be effective in selected COVID-19 patients. The objective is to present our experience of tocilizumab use in patients with severe COVID-19.

**Methods:** Observational retrospective cohort study. Hospitalized patients were evaluated by our multidisciplinary team for eventual use of tocilizumab. Patients with progressive ventilatory impairment and evidence of a hyperinflammatory state despite usual treatment received tocilizumab 8 mg/kg intravenous (maximum dose 800 mg), in addition to standard treatment. The use and time of use of mechanical ventilation (MV), the change of the Alveolar-arterial (A-a) gradient, of the ratio of arterial oxygen partial pressure to fractional inspired oxygen (PaO2/FiO2) and of inflammation laboratory parameters after 72 hours of tocilizumab use was evaluated.

**Results:** 29 patients received tocilizumab. 93.1% were men, 37.9% were obese, and 34.5% had hypertension. Of the 20 patients who were not on MV when receiving tocilizumab, 11 required non-invasive MV, for an average of five days, and one of them required intubation. A-a gradient, PaO2/FiO2, and inflammation parameters improved significantly. A better lymphocyte count, which improved significantly after tocilizumab use, was significantly associated with less use of MV. Five patients presented positive culture samples after tocilizumab, three being of clinical significance. A lower lymphocyte count was associated with having a positive culture. No other significant adverse events were seen.

**Conclusion:** Our study suggests the utility and shows the safety of tocilizumab use in COVID-19 patients who have respiratory failure and evidence of hyperinflammation. Lymphocyte improvement was a predictor of good response.

**Key-points:**

- The use of tocilizumab in patients with severe COVID-19 was safe.
- Most of the patients presented a good response in terms of ventilatory and inflammatory parameters.
- Lymphocyte improvement after using tocilizumab was the main predictor of a good outcome.

## Introduction

Coronavirus disease 2019 (COVID-19), caused by the severe acute respiratory syndrome coronavirus 2 (SARS-CoV-2), can lead to life-threatening respiratory symptoms[1].

The pathogenesis of the disease is complex and not yet fully understood[2, 3], however, as a consequence of lytic viral infection macrophage activation, and immune cell recruitment to alveoli, a significant increase in the secretion of proinflammatory cytokines occurs, referred to as “cytokine release syndrome” (CRS)[4, 5]. Different phases have been proposed in the course of this disease, stage I (mild) of early infection and active viral replication, stage II (moderate) of lung involvement without (IIa) or with hypoxia (IIb), where viral replication and local inflammation can coexist, and stage III (severe), with systemic hyperinflammation[6]. Several immunomodulatory strategies have been proposed to treat hyperinflammation in phases II and III and are currently being studied[7–11].

Interleukin-6 (IL-6) seems to have a pivotal role in the production of diffuse alveolar damage and acute respiratory distress syndrome[12, 13]. Thus, it is possible to conceive an IL-6 inhibition strategy to control CRS in patients with progressive hyperinflammation. Tocilizumab is a humanized anti IL-6 receptor antibody approved for the treatment of rheumatoid arthritis, juvenile idiopathic arthritis, giant cell arteritis, and life-threatening cytokine release induced by chimeric-antigen receptor T-cell therapy[14].

Faced with the possibility of not having the health resources to confront the pandemic, and in the absence of treatments with demonstrated efficacy, we constituted a multidisciplinary, composed of physicians from various specialties including rheumatology, immunology, hematology, infectious diseases, intensive care medicine and pulmonologists, with the aim of developing a guide to consider the use of tocilizumab, with a strict protocol to evaluate the patient and suggest its use for specific patients.

Our purpose is to present our experience of tocilizumab use in a cohort of patients with severe COVID-19, with emphasis on ventilatory results, inflammatory parameters, and adverse events.

## Methods

### Study design

Observational retrospective cohort study.

### Setting

Patients hospitalized in Clínica Alemana, Santiago, Chile, with a diagnosis of COVID-19, for whom evaluation was requested by our multidisciplinary team for eventual tocilizumab use. The results of the patients evaluated between April 13 and June 16, 2020 who received tocilizumab and who were discharged or died before July 18, 2020 are reported. All data was collected by this multidisciplinary team from the patient’s medical record by the multidisciplinary team.

### Participants

The multidisciplinary team evaluated each case by reviewing the patient’s medical file and discussing with the treating physician. Criteria were previously established to consider the use of tocilizumab: Inclusion criteria:

1. Age ≥18 years.
2. SARS-CoV-2 confirmed infection according to institutional criteria (polymerase chain reaction [PCR], antibodies, and/or compatible images).
3. ≥seven days from the onset of symptoms.
4. Evidence of severe COVID-19:
  a. At least two of the following:
    i. Respiratory rate>24x.
    ii. Multiple pulmonary infiltrates on chest radiography or computed axial tomography and/or increase in infiltrates.
    iii. O2 saturation <93% with a fraction of inspired oxygen (FiO2) of 21% or O2 requirement to maintain saturation>92%.
    iv. Fever>38°C.
  b. And at least two of the following:
    i. C-reactive protein (CRP) > 5mg/dL (or relative increase >50% in less than 48hrs).
    ii. Alveolar-Arterial (A-a) O2 gradient with deterioration>50%.
    iii. Increase ≥25% of ferritin with respect to the previous value within the last 48 hours or isolated value ≥2000 ng/mL.
    iv. Neutrophil-lymphocyte ratio (NLR)>3.12 or lymphopenia <700/mm3.
    v. Fibrinogen increase ≥25% or ≥800 mg/dL.

Exclusion criteria:

1. Not controlled infection by bacteria, fungi or non-COVID viruses.
2. Active tuberculosis.
3. Any concurrent immunomodulatory medication or intervention that the treating physician believes would put the patient at increased risk.
4. History of diverticulitis or intestinal perforation.
5. Absolute neutrophil count <1,000/mm3, platelets <50,000/mm3.
6. Known allergy to tocilizumab, or to any other anti-IL-6 agent.
7. Any condition that, in the opinion of the treating physician or the multidisciplinary group, would increase the patient’s risk when receiving tocilizumab.

If the patient met the criteria, the indication for tocilizumab was discussed within the multidisciplinary group. If tocilizumab use was recommended, this suggestion was communicated to the treating physician and to the patient or his/her family, explaining the risks and benefits, specifying that the use was off-label. If its use was accepted, tocilizumab was administered as a single 60 minutes intravenous drip infusion. The dose used was 8 mg/kg, with a maximum of 800 mg.

### Variables

The baseline characteristics of the patients were recorded. Data from clinical chart was recorded, specifying the level of care, the type of ventilatory support (nasal cannula, non-invasive mechanical ventilation [NIMV], invasive mechanical ventilation [IMV]), the use of prone position, hydroxychloroquine (HCQ), convalescent plasma, corticosteroids (CS), and tocilizumab. Several laboratory parameters were considered including lymphocyte and neutrophil counts, erythrocyte sedimentation rate (ESR), CRP, ferritin, D-dimer, fibrinogen, Lactate dehydrogenase (LDH), transaminases, and IL-6. The A-a gradient and the ratio of arterial oxygen partial pressure to fractional inspired oxygen (PaO2/FiO2) were calculated.

The change, between the time of tocilizumab use and 72 hours after, in laboratory and ventilatory parameters was analyzed.

The use and time of use of mechanical ventilation (MV), and the percentage improvement of the A-a gradient and of PaO2/FiO2 after 72 hours of tocilizumab use were defined as outcomes.

Deaths, non-covid-19 infections and events not attributable to the usual course of the disease were recorded and analyzed.

### Data sources/measurement

Data was obtained from the patient’s clinical record in the institutional electronic clinical file. The use of MV was evaluated as a qualitative and quantitative variable (days of use), and improvements in the A-a gradient and PaO2/FiO2 were evaluated quantitatively. Non-COVID-19 infections and adverse events not attributable to the usual course of the disease were evaluated as qualitative variables.

### Statistical methods

The data was analyzed using IBM SPSS Statistics for Mac OS, Version 24.0. Armonk, NY. Categorical variables were described as frequencies and/or percentages, and quantitative variables were described as mean and standard deviation (SD) for those normally distributed or median and interquartile range (IQR) for those not normally distributed. Changes in parameters were analyzed using T-test for paired variables or Wilcoxon signed-rank sum test, as appropriate. The associations between variables and outcomes were evaluated using linear or logistic regressions for univariate and multivariate analysis. The covariates included in the multivariate analysis model were age, sex, body mass index, diabetes, hypertension, asthma, obstructive sleep apnea, smoking status, days with symptoms. Statistical significance was defined as P<0.05. In case of missing data, the analysis was performed with the available data.

### Institutional Review Board

The Scientific Ethics Committee of the Clínica Alemana-Universidad del Desarrollo medicine faculty approved the study prior to data collection.

## Results

### Participants

29 of 68 evaluated patients (42.6%) received Tocilizumab.

### Descriptive data

Baseline characteristics: The mean age of the patients was 56.9 years, and only two were women. 37.9% were obese and the most frequent comorbidity was hypertension. All patients presented lymphopenia and elevated inflammatory markers (Table 1).

**Table 1.** Descriptive data.

| Table 1. Descriptive data |  |  |
| --- | --- | --- |
| Variable | Number of patients with data | Results |
| Age, mean (SD) | 29 | 56.9 (13.6) |
| Men, n (%) | 29 | 27 (93.1) |
| BMI, mean (SD) | 29 | 28.2 (4.5) |
| BMI ≥30, n (%) | 29 | 11 (37.9) |
| Smoking, n (%) | 29 | 5 (17.2) |
| Hypertension, n (%) | 29 | 10 (34.5) |
| DM, n (%) | 29 | 4 (13.8) |
| OSA, n (%) | 29 | 3 (10.3) |
| Asthma, n (%) | 29 | 1 (3.4) |
| Unit upon admission, n (%) | 29 |  |
| Basic |  | 18 (62.1) |
| Intermediate care |  | 11 (37.9) |
| Days with symptoms upon admission, median (IQR) | 29 | 8 (5-9) |
| Days of hospitalization, median (IQR) | 29 | 14 (12-17) |
| PaO<sub>2</sub>/FiO<sub>2</sub> (mmHg) on admission, mean (SD) | 28 | 303.3 (103.8) |
| Expected Alveolar-arterial gradient, median (IQR) | 29 | 14.7 (11.7-15.7) |
| A-a gradient (mmHg) on admission, median (IQR) | 28 | 40.9 (18.4-82.5) |
| Lymphocytes/mm<sup>3</sup> on admission, median (IQR) | 28 | 958 (783.5-1202) |
| NLR at admission, median (IQR) | 27 | 4.2 (3.4-7.3) |
| ESR (N 2-30 mm/hr) on admission, mean (SD) | 17 | 44.6 (29.8) |
| CRP (N 0.1-0.5 mg/dL) on admission, median (IQR) | 25 | 4.5 (2.3-11.1) |
| Ferritin (N 4.6-204 ng/mL) on admission, median (IQR) | 22 | 1454.2 (899.2-3230.2) |
| D-dimer (N 0-500 ng/mL) on admission, median (IQR) | 26 | 575 (359-819) |
| Troponin (N <14 ng/L) on admission, median (IQR) | 23 | 7.5 (4.2-13.8) |
| Fibrinogen (N 238-498 mg/dL) on admission, mean (SD) | 20 | 527.3 (141) |
| LDH (N 135-214 U/L) on admission, mean (SD) | 26 | 331.4 (128.4) |
| SD: standard deviation; BMI: body mass index; DM: diabetes mellitus; OSA: obstructive sleep apnea; IQR: interquartile range; PaO<sub>2</sub>/FiO<sub>2</sub>: ratio of arterial oxygen partial pressure to fractional inspired oxygen; A-a: Alveolar-arterial; NLR: Neutrophil-lymphocyte ratio; N: normal range; ESR: erythrocyte sedimentation rate; CRP: C-reactive protein; LDH: lactate dehydrogenase. |  |  |

Tocilizumab: Tocilizumab was used at a median of four days of hospitalization and 11 with symptoms. Of the 17 patients (58.6%) who received tocilizumab while in the basic hospitalization unit, 11 (64.7%) were subsequently transferred to intermediate care, and of those 11 patients, three (27.3%) required admission to intensive care. Of the eight patients who received the medication while at the intermediate care unit, two (25%) had to be admitted to intensive care afterwards. The other four patients received the medication while at the intensive care unit. The median cumulative dose of CS (equivalent to prednisone) at the time of tocilizumab use was 150 mg. IL-6 was elevated before tocilizumab use, and afterwards it was measured again in five patients (median 104; IQR 89.8-446) (Table 2).

**Table 2.** Tocilizumab use.

| **Table 2. Tocilizumab use** |  |  |
| --- | --- | --- |
| **Variable** | **Number of patients with data** | **Results** |
| Days with symptoms when used, median (IQR) | 29 | 11 (10-14) |
| Unit upon use, n (%) | 29 | 29 (100) |
| Basic | 29 | 17 (58.6) |
| Intermediate care | 29 | 8 (27.6) |
| Intensive care | 29 | 4 (13.8) |
| Prednisone equivalent dose (mg) of CS, accumulated before the use of tocilizumab, median (IQR) | 29 | 150 (100-300) |
| IL-6 (N 0-3.4 pg/ml) at the time of tocilizumab, median (IQR) | 22 | 35.2 (16.1-85.1) |
| Days of hospitalization after receiving tocilizumab, median (IQR) | 29 | 10 (8-13) |
| Days in intermediate or intensive care after receiving tocilizumab, median (IQR) | 29 | 6 (3-9) |
| Tocilizumab administered to a patient on MV*, n (%) | 29 | 9 (31) |
| Days on MV, mean (SD) | 9 | 8.6 (4.4) |
| Days on MV after tocilizumab, mean (SD) | 9 | 7.3 (4.3) |
| Patients, excluding those already on MV, that required MV* after tocilizumab, n (%) | 20 | 11 (55) |
| Days on MV, mean (SD) | 11 | 5.5 (3.7) |
| Lymphocyte count % improvement from the day of tocilizumab use until 72 hours later, median (IQR) | 29 | 92.1 (41.4-183.2) |
| A-a gradient % improvement from the day of tocilizumab use until 72 hours later, median (IQR) | 25 | 30.7 (13.9-53.8) |
| PaO<sub>2</sub>/FiO<sub>2</sub> % improvement from the day of tocilizumab use until 72 hours later, median (IQR) | 25 | 23 (8.8-58.4) |
| *Including non-invasive and invasive MV.<br>IQR: interquartile range; CS: corticosteroids; IL-6: interleukin-6; N: normal range; MV: mechanical ventilation; A-a: Alveolar-arterial; PaO<sub>2</sub>/FiO<sub>2</sub>: ratio of arterial oxygen partial pressure to fractional inspired oxygen. |  |  |

Other interventions: (Table 3) six patients received HCQ (400 mg every 12 hours the first day and then 200 mg every 12 hours for four days, five days in total).

**Table 3.** Non-tocilizumab interventions.

| **Table 3. Non-tocilizumab interventions** |  |  |
| --- | --- | --- |
| **Variable** | **Number of patients with data** | **Results** |
| Use of CS, n (%) | 29 | 28 (96.6) |
| Days of use, median (IQR) | 29 | 9 (7-13) |
| Prednisone equivalent dose (mg), accumulated during hospitalization, mean (SD) | 29 | 725.9 (251.7) |
| Hydroxychloroquine use, n (%) | 29 | 6 (20.7) |
| Use of prone position | 29 | 24 (82.8) |
| Use of convalescent plasma, n (%) | 29 | 8 (27.6) |
| Days with symptoms when used, mean (SD) | 8 | 6.9 (1.5) |
| Maximum ventilatory support during hospitalization, n (%) | 29 |  |
| NC |  | 8 (27.6) |
| HFNC |  | 1 (3.4) |
| Non-invasive MV |  | 16 (55.2) |
| Invasive MV |  | 4 (13.8) |
| CS: corticosteroids; IQR: interquartile range; SD: standard deviation; NC: nasal canulla; HFNC: high flow nasal canulla; MV: mechanical ventilation |  |  |

### Outcome data

Laboratory parameters: From admission to the day of tocilizumab use, lymphocytes decreased by a mean of 30.7% (SD 36). After 72 hours of using tocilizumab there was a significant improvement in the lymphocyte count, NLR, ESR, CPR, and fibrinogen. A mild and transient, but statistically significant, rise in alanine aminotransferase, gamma-glutamyl transferase and D-dimer was observed (Table 4).

**Table 4.** Changes in laboratory and ventilatory parameters after using tocilizumab.

|  |  |  |  |  |
| --- | --- | --- | --- | --- |
| **Variable** | **Number of patients with data** | **At the time of receiving tocilizumab** | **After 72 hours of receiving tocilizumab** | **p** |
| *PaO<sub>2</sub>/FiO<sub>2</sub>, mean (SD)* | 25 | 219.5 (56.4) | 262.1 (79.8) | 0.047 |
| *A-a gradient, median (IQR)* | 25 | 110.2 (91.1-144.5) | 74.2 (50-102.1) | 0.007 |
| *Lymphocytes/mm<sup>3</sup>, median (IQR)* | 29 | 571 (473-797) | 1266 (792-1534) | <0.001 |
| *NLR, median (IQR)* | 29 | 13.8 (10-20.3) | 5.6 (3.2-7.3) | <0.001 |
| *ESR (N 2-30 mm/hr), median (IQR)* | 26 | 61 (47-80) | 39 (30-52) | <0.001 |
| *CRP (N 0.1-0.5 mg/dL), median (IQR)* | 29 | 13.1 (8.7-20.3) | 1.4 (0.8-2.3) | <0.001 |
| *Ferritin (N 4.6-204 ng/mL), median (IQR)* | 24 | 2346.7 (1800.9-3818.9) | 2434.9 (1248.1-4769.4) | NS |
| *D-dimer (N 0-500 ng/mL), median (IQR)* | 23 | 582 (354.5-957.5) | 1028 (582.5-1527) | 0.001 |
| *Fibrinogen(N238-498 mg/dL), median (IQR)* | 20 | 648 (521-718.5) | 391.5 (338.5-445.5) | <0.001 |
| *AST (N 5-32 U/L), median (IQR)* | 26 | 38.5 (29-58) | 59 (29-91) | NS |
| *ALT (N 5-33 U/L), median (IQR)* | 26 | 45 (26-67) | 97 (56-174) | 0.003 |
| *GGT (N 3-40 U/L), median (IQR)* | 26 | 105.5 (47-212) | 174 (103-366) | <0.001 |
| *LDH (N 135-214 U/L), mean (SD)* | 26 | 344.3 (97.4) | 341.4 (110.5) | NS |
| PaO<sub>2</sub>/FiO<sub>2</sub>: ratio of arterial oxygen partial pressure to fractional inspired oxygen; A-a: Alveolar-arterial; NLR: Neutrophil-lymphocyte ratio; N: normal range; ESR: erythrocyte sedimentation rate; N: normal range; CRP: C-reactive protein; AST: Aspartate aminotransferase; ALT: Alanine aminotransferase; GGT: Gamma-glutamyl transferase; LDH: lactate dehydrogenase. |  |  |  |  |

Ventilatory parameters: The improvement in PaO2/FiO2 (23%) and A-a gradient (30.7%) after 72 hours of tocilizumab use was statistically significant. Nine patients received tocilizumab while on MV. Of the remaining 20 patients, 11 (55%) required MV after using tocilizumab. Of these 11 only one required IMV (the other three patients who used IMV required it before tocilizumab use) (Tables 2, 4).

Infections: Seven culture samples were positive after tocilizumab use, in five patients (17.2%). Only three of them presented a proven clinical infection (Table 5).

**Table 5.**
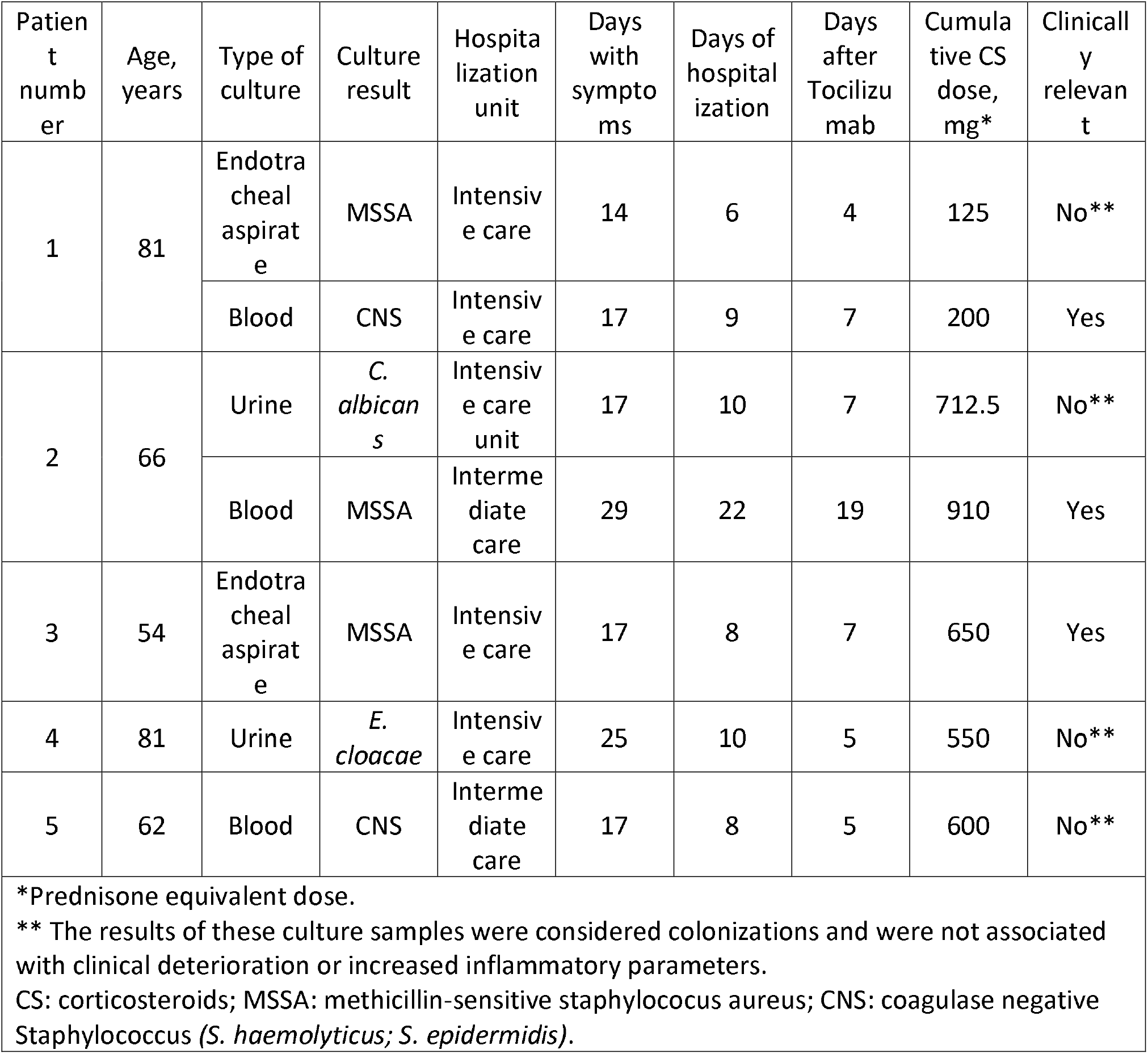
Positive cultures.

| Patient number | Age, years | Type of culture | Culture result | Hospitalization unit | Days with symptoms | Days of hospitalization | Days after Tocilizumab | Cumulative CS dose, mg* | Clinically relevant |
| --- | --- | --- | --- | --- | --- | --- | --- | --- | --- |
| 1 | 81 | Endotracheal aspirate | MSSA | Intensive care | 14 | 6 | 4 | 125 | No** |
|  |  | Blood | CNS | Intensive care | 17 | 9 | 7 | 200 | Yes |
| 2 | 66 | Urine | *C. albicans* | Intensive care unit | 17 | 10 | 7 | 712.5 | No** |
|  |  | Blood | MSSA | Intermediate care | 29 | 22 | 19 | 910 | Yes |
| 3 | 54 | Endotracheal aspirate | MSSA | Intensive care | 17 | 8 | 7 | 650 | Yes |
| 4 | 81 | Urine | *E. cloacae* | Intensive care | 25 | 10 | 5 | 550 | No** |
| 5 | 62 | Blood | CNS | Intermediate care | 17 | 8 | 5 | 600 | No** |

\*Prednisone equivalent dose.
\*\* The results of these culture samples were considered colonizations and were not associated with clinical deterioration or increased inflammatory parameters.
CS: corticosteroids; MSSA: methicillin-sensitive staphylococcus aureus; CNS: coagulase negative Staphylococcus (*S. haemolyticus*; *S. epidermidis*).

The first patient was an 81-year-old man, with history of rheumatoid arthritis and chronic CS use, who received tocilizumab on the second day of hospitalization while on NIMV (with a procalcitonin [PCT] of 0.28 ng/mL), was intubated on the third day, and on the sixth day a methicillin-sensitive *Staphylococcus aureus* was identified in the endotracheal aspirate culture, without clinical impact. On the ninth day of hospitalization a methicillin-resistant *Staphylococcus epidermidis* was identified in two blood and one catheter tip samples. This was compatible with a central line associated bacteremia and the central line was removed. The fever disappeared without additional antibiotic treatment. He was discharged after 29 days of hospitalization.

The second patient was a 66-year-old man with a urine culture positive for candida albicans (>100,000 colony forming units) with non-inflammatory urine (PCT of 0.45 ng/mL at the time of tocilizumab use). This was an asymptomatic candiduria. Twelve days later he presented two positive blood cultures for methicillin-sensitive Staphylococcus aureus. He had an elbow bursitis and a retroperitoneal hematoma at the same time. He was successfully treated with intravenous cefazolin and was discharged after 44 days of hospitalization.

The third patient was a 54-year-old man, who presented a positive culture of the endotracheal aspirate for a methicillin-sensitive Staphylococcus aureus. The bronchoalveolar lavage culture of the same day was negative. He was successfully treated with intravenous antibiotics.

In all patients the PCT measured on the day of tocilizumab use had a median value of 0.17 ng/mL (IQR 0.1-0.38; <0.5 ng/mL= low risk of sepsis). In the five patients who presented a positive culture, PCT had a median value of 0.26 (IQR 0.17-0.28), at the time of tocilizumab use. There was no statistically significant difference in the PCT between these two groups.

Thus, three patients presented a clinically relevant infection, and none of these had severe course.

Adverse events: Events out of the expected course for COVID-19, after receiving tocilizumab, were described in 6 male patients (20.7%).

One patient presented leukopenia of 2,800/mm3 with neutropenia of 1,425/mm3, which normalized ten days after discharge. Two patients had mild elevation of transaminases. The study of known hepatotropic agents was negative, thus a direct effect of SARS-CoV-2 infection or drugs was considered as possible causes. One of these patients had already had his post-discharge evaluation and transaminases had normalized.

On the 27th day of hospitalization, an 81-year-old patient (patient number 1 in table 5) who required intubation presented a small left para-aortic pleural collection, possibly with blood content, on computed axial tomography. This did not mean any change for the course of his hospitalization, and he was discharged after 29 days of hospitalization.

Two patients presented pulmonary thromboembolism. The first case occurred in a 54-year-old patient, on his eighth day of hospitalization and on his seventeenth day of symptoms, seven days after receiving tocilizumab and after three days connected to IMV, being on prophylactic anticoagulation, without D-dimer level of that day. Pulmonary thromboembolism was bilateral and difficult to anticoagulate, and his hospitalization lasted 26 days, but he was discharged in good condition. The second patient was 66 years old and presented a left basal subsegmental pulmonary thromboembolism, on day 17 of hospitalization and on the 25th day of symptoms, 14 days after receiving tocilizumab and after 15 days on MV (nine of them on IMV), being on prophylactic anticoagulation, with a D-dimer of 1.751 ng/mL (11 days before it was 5.858 ng/mL). After seven days of anticoagulation, the patient presented an extensive retroperitoneal hematoma. He was discharged after 36 days of hospitalization in good condition.

The only variable that was significantly associated with presenting pulmonary thromboembolism was the number of days of hospitalization at the intermediate or intensive care unit (Exp (B) 1.332; 95% C.I. 1.004-1.765; p=0.46).

Deaths: Two patients died (6.9%). The first patient was an 81-year-old man with history of prostate cancer successfully treated surgically. The patient required IMV after 1 day of hospitalization and received tocilizumab in this condition on the fifth day of hospitalization and twentieth of symptoms. The patient managed to transition to NIMV temporarily. The only positive culture was a urine culture with *Enterobacter cloacae* (patient number 4 in table 5). The patient followed a progressive course with respiratory and hemodynamic deterioration, with elevated levels of troponin and N-terminal pro brain natriuretic peptide, and presented a sharp drop in his hematocrit (from 35% to 23%, being anticoagulated with dalteparin) before dying in the eleventh day of hospitalization.

The second patient was a 73-year-old man with history of obstructive sleep apnea and possible interstitial lung disease. He received tocilizumab on the third day of hospitalization and eighth of symptoms while on NIMV. He did not present notable infections or events, but had progressive respiratory deterioration, and after 7 days in NIMV it was decided to limit therapeutic efforts and the patient was not intubated. He died on the twelfth day of hospitalization.

No significant associations were found between the different variables and death.

### Main results

Lymphocyte count: A greater percentage improvement in the lymphocyte count between the day of tocilizumab use and 72 hours afterwards was significantly associated with a higher number of days with symptoms when tocilizumab was used, and a higher cumulative dose of corticosteroids. These associations did not maintain statistical significance in the multivariate analysis (Appendix Table 1).

Ventilatory parameters: A greater percentage improvement in the A-a gradient was significantly associated with a lower level of D-dimer when receiving tocilizumab, and a greater improvement in PaO2/FiO2 was associated with a higher level of ESR and lower D-dimer at the time of receiving tocilizumab. These associations were weak, and none remained significant after multivariate analysis (Appendix Table 1). Mechanical ventilation: A lower lymphocyte count 72 hours after tocilizumab use was associated with requiring MV in patients who were not on MV at the time of receiving tocilizumab (table 6). In the 20 patients who received tocilizumab without using MV, there were no significant differences between the 11 patients who required MV later and the 9 who did not, except in the lymphocyte count 72 hours after receiving tocilizumab (p = 0.004) and their percentage of improvement (p = 0.016). At the time of receiving tocilizumab, the median number of lymphocytes in those who subsequently required MV was 682 (IQR 484-767) and rose to 807 (IQR 666-1335) 72 hours after tocilizumab, a median improvement of 51.7% (IQR 30.3-79.3). In those who did not require MV after tocilizumab, the lymphocytes rose from 737 (IQR 571-806) to 1602 (IQR 1402-2087), a median improvement of 157.8% (IQR 99.7-180.6). A greater number of days in MV after tocilizumab was significantly associated with worse PaO2/FiO2 and A-a gradient, and with lower lymphocyte count, but only the A-a gradient and the lymphocyte count maintained statistical significance in the multivariate analysis (Appendix Table 1). Furthermore, a lower number of lymphocytes on admission was associated with requiring IMV (table 6).

**Table 6.** Logistic regression.

| Table 6. Logistic regression |  |  |  |  |
| --- | --- | --- | --- | --- |
| Outcome | Variable* | Exp(B) | 95% C.I. Exp(B) | p |
| ***MV use, excluding patients already on MV when tocilizumab was administrated*** | Lymphocytes 72 hours after tocilizumab | 0.996 | 0.993-1 | 0.032 |
| ***Invasive MV*** | Lymphocytes on admission | 0.995 | 0.99-1 | 0.032 |
| **Positive culture** | Age | 1.111 | 1-1.234 | 0.05 |
|  | Lymphocytes on admission | 0.996 | 0.991-1 | 0.038 |
|  | Lymphocytes at the time of receiving tocilizumab | 0.993 | 0.986-1 | 0.039 |
|  | NLR 72 hours after Tocilizumab | 1.570 | 1.025-2.405 | 0.038 |
| *No association was significant in the multivariate model.<br>MV: mechanical ventilation; NLR: Neutrophil-lymphocyte ratio. |  |  |  |  |

Infections: Older age, low lymphocyte count and a high NLR were associated with a greater possibility of presenting a positive culture (Table 6).

Deaths: No associations were found.

## Discussion

This is the first Latin American report to evaluate the use of tocilizumab in patients hospitalized for severe COVID-19.

Our cohort corresponds to relatively young Latin American patients, mainly men, with a significant prevalence of comorbidities known to be strong risk factors for severe COVID-19[15]. The patients presented a severe disease and 69% required MV. All patients presented biomarkers compatible with hyperinflammation, impaired gas exchange, and rapid worsening of these despite receiving usual treatment. The use of hydroxychloroquine was infrequent since its prescription decreased when reports were published that warned of the ineffectiveness and risk of adverse events of its use[16].

Rapid clinical deterioration in most of our patients, in a phase of hyperinflammatory disease, led to the indication of immunomodulatory therapy, at first with CS. In practice, we used a sequential strategy in which the use of tocilizumab was decided only once despite the use of corticosteroids a worsening of the patient condition was observed. CS usefulness in COVID-19 remains controversial[17], however, in a recently published trial, the use of dexamethasone (6 mg every day for a median six days, approximately a cumulative dose equivalent to 225 mg of prednisone) resulted in lower 28-day mortality[18]. In our cohort the cumulative dose of CS, equivalent to prednisone, was higher, with a median of 725.9 mg. A recently published retrospective analysis also showed good results with a high dose of CS and subsequent use of tocilizumab[19]. Undoubtedly, the routine CS use with a high cumulative dose results in an important confounding factor when interpreting efficacy and safety data.

In our patients, 72 hours after tocilizumab use, a dramatic improvement was observed in ventilatory parameters and in most CRS biomarkers. D-dimer was highly variable from day to day in our patients. Although its values increased 72 hours after tocilizumab and seemed to be associated with improvements in A-a gradient and PaO2/Fio2, the changes in its levels over the course of the hospitalization make it difficult to draw conclusions about its associations and predictive role.

A higher lymphocyte count was associated with less use of MV. Lymphopenia is a common finding in severe COVID-19, with almost all subsets affected, and it has been suggested as a poor prognostic factor and reliable indicator of severity and mortality risk[20–22]. It is not clear if SARS-CoV-2 directly affects T cells or if the triggered cytokine release drives depletion and exhaustion of T cells[23]and impairs the cytotoxic activity of CD8 T cells and NK cells, which are crucial for antiviral immunity[24]. Strategies to improve T cell counts and function may be critical to reduce disease severity. It is expected that the use of CS would be associated with greater lymphopenia and worse NLR, but what we observed was that after the use of tocilizumab these parameters improved significantly. It can be hypothesized that blocking IL-6 could help in COVID-19 not only by inhibiting directly the exaggerated inflammatory response, but also by restoring cellular immunity which is crucial for virus clearance.

After multivariate analysis, the decrease in the A-a gradient, but not the increase of PaO2/FiO2, correlated with a lower number of days of MV use after tocilizumab infusion. This is because the gradient is a more sensitive index to evaluate disturbances in gas exchanges[25].

Assessing the safety of tocilizumab use in a COVID-19 patient is of the utmost importance. Despite the increase in transaminase values after tocilizumab use in our patients, this was transitory and not associated with liver failure. Two patients (6.9%) suffered from pulmonary thromboembolism, which seems to be within the reported rates for severe COVID-19[26].

In our experience, infections are the most important factor influencing fear of tocilizumab use in COVID-19 patients. Five patients of our cohort had a positive blood or urine culture, all of them hospitalized more than seven days and in intermediate or intensive care units when presenting the positive culture. Only three patients presented a clinically relevant infection, and none of these presented a severe course. Lower lymphocyte count and worse NLR were associated with a greater probability of having a positive culture. More than an immunosuppressive agent, tocilizumab acts as an immunomodulator, and our patients significantly raised their lymphocytes after its use, which was associated with a lower risk of infection.

Two of our patients died (6.9%). The mortality rate in our center, as of July 16, 2020, counting 1076 hospitalized patients, is 5.3%. The rate of our cohort is slightly higher, but it should be considered that it consists of patients selected because of their severity.

Our study has several limitations. It is an observational study without control group. It represents a small number of patients from a single center. In the analysis of the results, it was not possible to have some data in all the patients. This report may not capture adverse events that may occur in the long term.

Our study is relevant because it represents the experience of a Latin American country, with a population poorly represented in other series. In addition, we provide more information to assist in choosing the best “window of opportunity” for tocilizumab use, specially the work of a multidisciplinary team that evaluated each patient in detail. Our team analyzed the pathophysiological rationality and the accumulated evidence and established criteria to consider the use of tocilizumab that considered the presence of an uncontrolled inflammatory response. Even when the patients met the established criteria, there was a group discussion weighing the risks and possible benefits before suggesting the use of the drug. Furthermore, we describe the use of tocilizumab together with high doses of CS, we detail the infections, adverse events, and deaths, and we analyze the variables associated with a better hospital stay. The provided data is useful to improve patient selection, trying to minimize risks and optimize efficacy.

Our study suggests the utility and shows the safety of tocilizumab use in COVID-19 patients who have respiratory failure and evidence of CRS. Lymphocyte improvement after using tocilizumab was the best predictor of good response to tocilizumab.

## Data Availability

All data referred to in the manuscript is available on request

## Declarations

### Authors’ contributions

All authors contributed equally with literature search, study design, data collection, data interpretation, and writing. S. Ibáñez perfomed the data analysis.

---

**Appendix Table 1.** Linear regressions.

| Appendix Table 1. Linear regressions |  |  |  |  |  |
| --- | --- | --- | --- | --- | --- |
| Outcome | Number of patients | Variable | R | Adjusted R<sup>2</sup> | P |
| *Lymphocyte improvement (%) from the day of tocilizumab use until 72 hours later* | 29 | Number of days with symptoms when receiving tocilizumab | 0.425 | 0.151 | 0.021 |
|  |  | Cumulative dose of corticosteroids at the time of receiving tocilizumab | 0.533 | 0.257 | 0.003 |
|  |  | Cumulative dose of corticosteroids 72 hours after the use of tocilizumab | 0.396 | 0.126 | 0.033 |
| *A-a gradient improvement (%) from the day of tocilizumab use until 72 hours later* | 25 | D-dimer at the time of receiving Tocilizumab | 0.436 | 0.190 | 0.033 |
| *PaO2/FiO2 improvement (%) from the day of tocilizumab use until 72 hours later* | 25 | ESR at the time of receiving Tocilizumab | 0.467 | 0.181 | 0.025 |
|  |  | D-dimer at the time of receiving Tocilizumab | 0.459 | 0.175 | 0.024 |
| *Number of days in MV after Tocilizumab* | 29 | Tocilizumab administered to a patient on MV | 0.46 | 0.183 | 0.012 |
|  |  | PaO2/FiO2 72 hours after Tocilizumab | 0.549 | 0.271 | 0.004 |
|  |  | A-a gradient at the time of receiving Tocilizumab* | 0.439 | 0.161 | 0.02 |
|  |  | A-a gradient 72 hours after Tocilizumab | 0.532 | 0.252 | 0.006 |
|  |  | A-a gradient improvement (%) from the day of tocilizumab use until 72 hours later | 0.425 | 0.145 | 0.034 |
|  |  | Lymphocytes 72 hours after tocilizumab* | 0.489 | 0.211 | 0.007 |
|  |  | NLR at the time of receiving Tocilizumab | 0.381 | 0.114 | 0.041 |
|  |  | NLR 72 hours after Tocilizumab | 0.419 | 0.145 | 0.024 |
| ***Number of days in MV, excluding patients already on MV when tocilizumab was administrated*** | ***20*** | PaO2/FiO2 72 hours after tocilizumab | 0.683 | 0.429 | 0.004 |
|  |  | PaO2/FiO2 improvement (%) | 0.512 | 0.209 | 0.043 |
|  |  | A-a gradient 72 hours after Tocilizumab | 0.7 | 0.454 | 0.003 |
|  |  | A-a gradient improvement (%) | 0.634 | 0.359 | 0.008 |
|  |  | Lymphocytes 72 hours after tocilizumab* | 0.628 | 0.361 | 0.003 |
|  |  | Lymphocyte improvement (%) | 0.469 | 0.176 | 0.037 |
| **\* Maintained statistical significance in multivariate analysis.**<br>**A-a: Alveolar-arterial; PaO2/FiO2: ratio of arterial oxygen partial pressure to fractional inspired oxygen; ESR: erythrocyte sedimentation rate; MV: mechanical ventilation; NLR: Neutrophil-lymphocyte ratio.** |  |  |  |  |  |

